# Identification of commensal gut microbiota signatures as predictors of clinical severity and disease progression in multiple sclerosis

**DOI:** 10.1101/2023.06.26.23291875

**Authors:** Theresa L Montgomery, Qin Wang, Ali Mirza, Deanna Dwyer, Qi Wu, Catherine A Dowling, Jacob WS Martens, Jennifer Yang, Dimitry N Krementsov, Yang Mao-Draayer

**Author notes:** Corresponding Authors: Dimitry Krementsov, PhD, Associate Professor, Department of Biomedical and Health Sciences, University of Vermont, USA, Yang Mao-Draayer, MD, PhD, Professor in Neurology, Autoimmunity Center of Excellence, University of Michigan, USA, Michigan Institute for Neurological Disorders.

## Abstract

**Background:** Multiple sclerosis (MS) is a chronic autoimmune disease of the central nervous system and a leading cause of neurological disability in young adults. Clinical presentation and disease course are highly heterogeneous. Typically, disease progression occurs over time and is characterized by the gradual accumulation of disability. The risk of developing MS is driven by complex interactions between genetic and environmental factors, including the gut microbiome. How the commensal gut microbiota impacts disease severity and progression over time remains unknown.

**Methods:** In a longitudinal study, disability status and associated clinical features in 60 MS patients were tracked over 4.2 ± 0.97 years, and the baseline fecal gut microbiome was characterized via 16S amplicon sequencing. Progressor status, defined as patients with an increase in Expanded Disability Status Scale (EDSS), were correlated with features of the gut microbiome to determine candidate microbiota associated with risk of MS disease progression.

**Results:** We found no overt differences in microbial community diversity and overall structure between MS patients exhibiting disease progression and non-progressors. However, a total of 45 bacterial species were associated with worsening disease, including a marked depletion in *Akkermansia*, *Lachnospiraceae,* and *Oscillospiraceae*, with an expansion of *Alloprevotella*, *Prevotella-9*, and *Rhodospirillales*. Analysis of the metabolic potential of the inferred metagenome from taxa associated with progression revealed a significant enrichment in oxidative stress-inducing aerobic respiration at the expense of microbial vitamin K_2_ production (linked to *Akkermansia*), and a depletion in SCFA metabolism (linked to *Lachnospiraceae* and *Oscillospiraceae*). Further, statistical modeling demonstrated that microbiota composition and clinical features were sufficient to robustly predict disease progression. Additionally, we found that constipation, a frequent gastrointestinal comorbidity among MS patients, exhibited a divergent microbial signature compared with progressor status.

**Conclusions:** These results demonstrate the utility of the gut microbiome for predicting disease progression in MS. Further, analysis of the inferred metagenome revealed that oxidative stress, vitamin K_2_ and SCFAs are associated with progression.

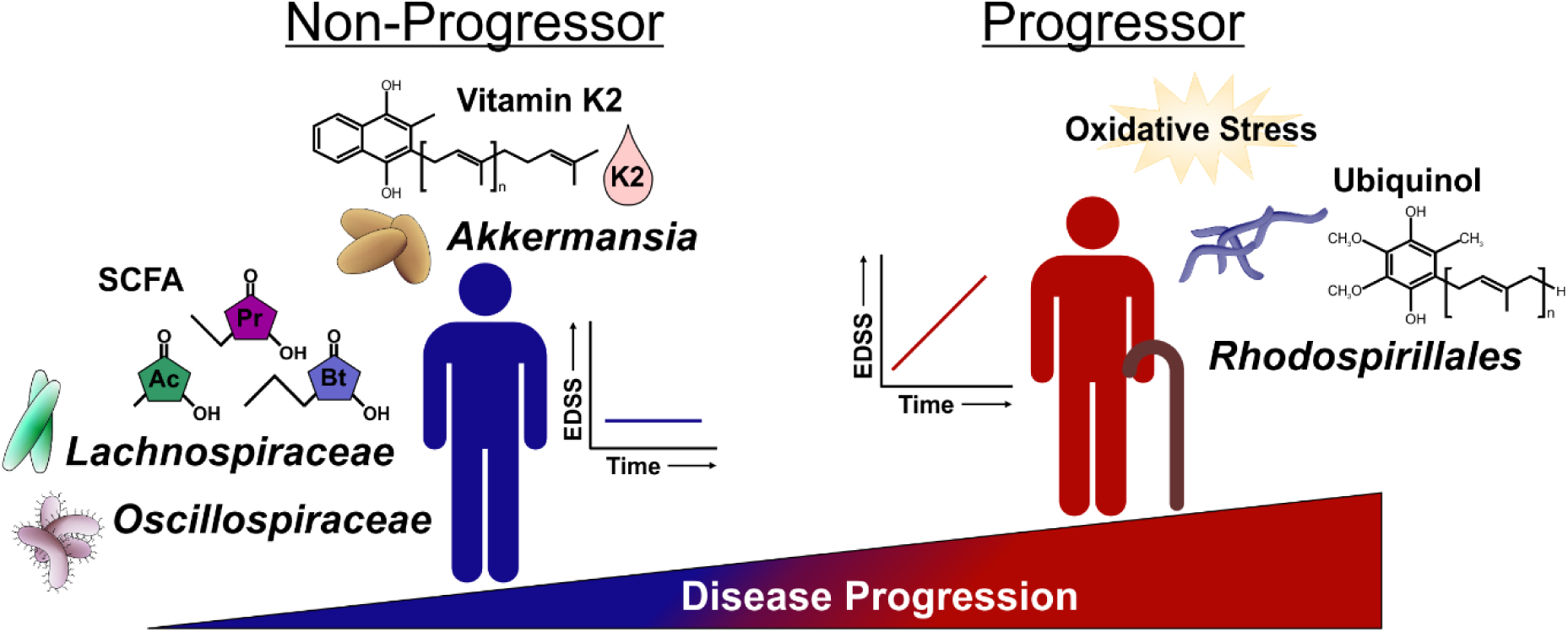

## BACKGROUND

Multiple sclerosis (MS) is a chronic demyelinating autoimmune disease of the central nervous system (CNS) that predominantly affects young adults [1–3]. However, the clinical presentation, onset, and disease course of MS may vary depending on genetic susceptibility and environmental factors, including the gut microbiome [4–7]. The severity and progression of MS is heterogeneous and varies from person to person [8]. Over time most cases of relapsing-remitting MS (RRMS) progressively worsen in severity and transition to secondary progressive MS (SPMS) with accumulation of disability [9]. Approximately 5-10% of patients experience only mild relapse without significant disease progression. These patients, classified as non-progressive benign MS (BMS/NPMS), never suffer from significant disabilities [10–12]. In recent years, the treatment paradigm for MS has shifted towards early aggressive intervention to prevent disease progression and irreversible CNS damage. Given that not all MS patients progress in their disease, there is an unmet need to identify patients who are at high risk for progression and require more aggressive treatment strategies to prevent long-term disability. Our prior studies identified candidate plasma biomarkers capable of distinguishing different progressive stages of MS [13]. Here, we examine the potential of using the gut microbiome as a prognostic approach to assess risk of progression.

Prior animal studies have shown that the gut microbiome can modulate peripheral and central nervous system immunity [14, 15]. Specifically, microbiota can alter demyelination and the permeability of the blood-brain barrier (BBB), which suggests potential as a predictive factor for relapse risk in MS [16–19]. Previous studies have also shown that MS patients have a distinct gut microbiome as compared to healthy patients, albeit with significant cohort to cohort variation, with the most consistent finding being increased abundance of *Akkermansia muciniphila* (*A. muciniphila*) in MS patients [18, 20, 21]. Functionally, several studies have suggested MS patient gut microbiota has a diminished capacity to produce short-chain fatty acids (SCFAs), which have a known role in limiting the production of pro-inflammatory cytokines [17, 22–27] and enhance the production of anti-inflammatory cytokines via an increase in regulatory T cells through SCFA G-protein-coupled receptors [28]. A decrease in abundance of SCFA-producing obligate anaerobes has been demonstrated in instances of increased oxygen-dependent aerobic respiration in the gut [29, 30]. A decrease in SCFAs in patients with SPMS has also been reported, demonstrating its possible utility as a marker for progression. Importantly, gastrointestinal symptoms are frequently reported in MS and often postulated to confound gut microbiome studies in these patients, especially in studies comparing healthy controls who typically lack constipation [31–35], yet no studies have directly documented the effects of this confounder on gut microbiota in MS patients.

While cross-sectional studies of the gut microbiome in MS have revealed some differences from healthy controls and associations with current disease activity [18, 27, 36–43], longitudinal studies exploring gut microbiota composition and clinical measures in MS remain sparse. Further, available longitudinal studies mostly focus on the potential relationship between the gut microbiome and relapse activity. Thus, it remains unclear whether gut microbiota are associated with MS progression and disability worsening. We utilized longitudinal tracking of disease severity (via the change in Expanded Disability Status Scale (EDSS)score) and the fecal gut microbiome (via 16S rRNA gene sequencing) to elucidate any relationship with MS progression. We took a novel approach comparing progressors vs. non-progressors across the spectrum of MS disease types. We found a total of 45 bacterial taxa were associated with worsening disease, including a marked depletion in *Akkermansia* and SCFA-producing *Lachnospiraceae* and *Oscillospiraceae* species, with an expansion of *Alloprevotella*, *Prevotella-9*, and *Rhodospirillales*. Pathway analysis on the inferred metagenome of taxa associated with progression revealed a significant enrichment in oxidative stress-inducing aerobic respiration at the expense of microbial vitamin K2 production, and a decrease in SCFA metabolism. Our study showed gut microbial signatures could also be used as potential predictors for disease progression across MS disease types. Gut microbiota aided in clustering of clinical features and identification of potential confounding effects of MS-associated symptoms; constipation was not a significant confounder when examining the gut microbiome in relationship to MS disease progression. Our results offer first insights into pathological mechanisms unique to progression, including specific microbial pathways, metabolites, or interactions that could contribute to worsening disease.

## RESULTS

### Gut microbiome baseline diversity and community structure does not differ in MS patients that exhibit disease progression

Fecal samples were collected from 60 MS patients recruited from the University of Michigan Medical School to profile the gut microbiome. In addition, initial disability status was assessed, followed by longitudinal tracking of EDSS from baseline for an average 4.2 ± 0.97 years (**Fig. 1A**). All subjects completed a clinical survey reporting subject demographics, disease subtype and history, medications, and any comorbidities (**Fig. 1A** and **Table 1**). In total, 13 patients (21.7%) had benign MS (BMS), 32 (53.3%) relapsing-remitting MS (RRMS), 10 (16.7%) secondary-progressive MS (SPMS), and 5 (8.3%) were diagnosed with primary-progressive MS (PPMS)

**Figure 1.**
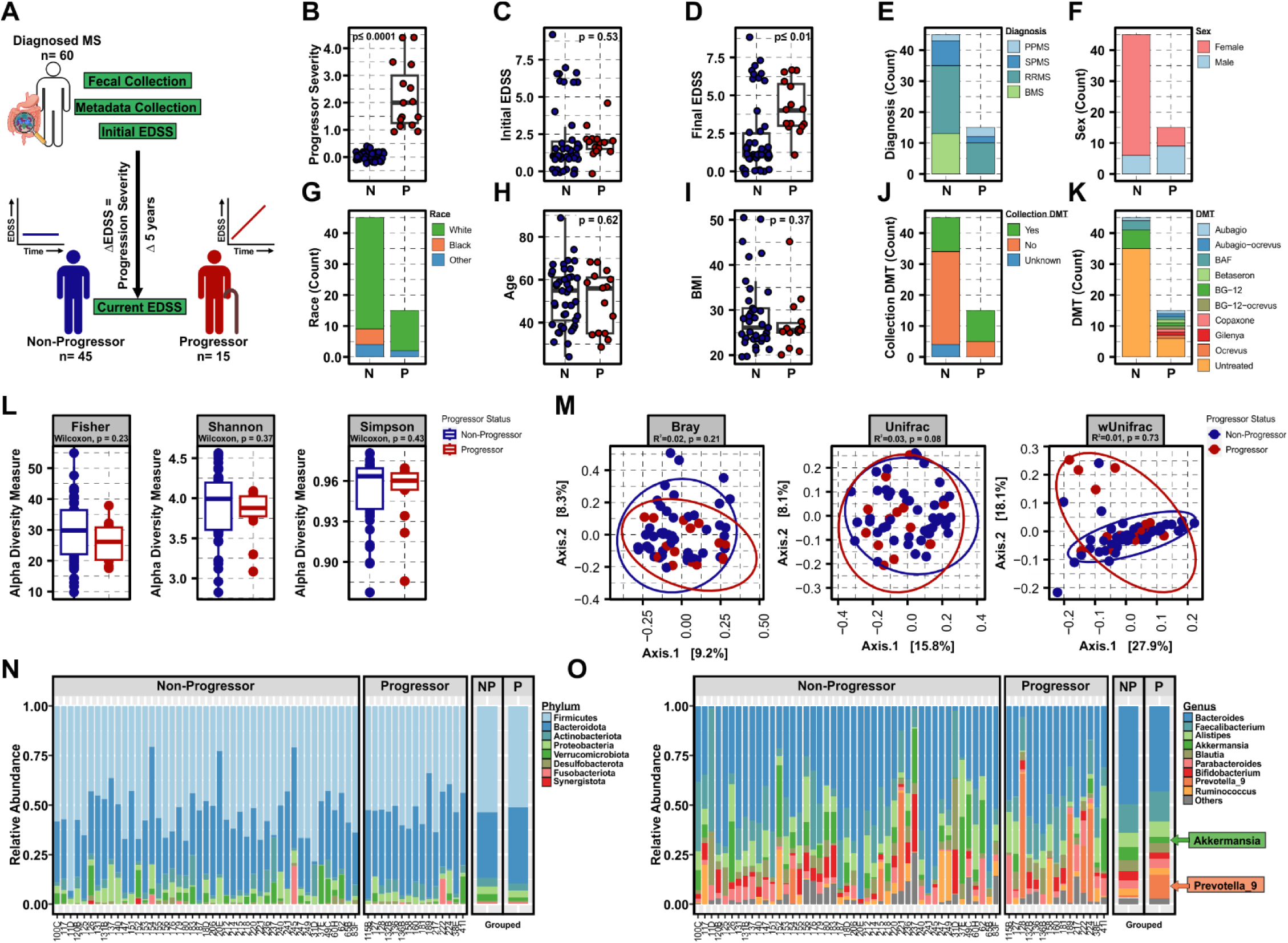
MS disease progression is not associated with differences in baseline gut microbiome alpha and beta diversity. (**A**) Schematic of clinical study design depicting participant binning strategy based on longitudinal disease progression where a non-progressor = change in EDSS of ≤|0.5| and a progressor = change in >|0.5|. Bar graphs and scatter plots of selected study metadata as outlined in **Table 1**, segregated by participant progressor status, including: (**B**) progressor severity (change in EDSS over study period), (**C**) initial EDSS, (**D**) final EDSS, (**E**) MS-subtype, (**F**) sex (**G**) self-identified race, (**H**) age, (**I**) BMI, (**J**) collection DMT status (**K**) DMT prior history. p-values represent a two-tailed binomial test of significant deviation from expected outcome of a 50% division of between progressors and non-progressors for categorical data, or standard t-test for numerical data, with significance at p≤0.05. Complete statistical analysis of subject metadata is included in **Table 1** and **2**. (**L**) Alpha (Fisher, Shannon and Simpson) and (**M**) beta (Bray-Curtis dissimilarity, Unifrac, wUnifrac) diversity analysis in progressors and non-progressors, as analyzed using Wilcoxon rank sum non-parametric or Adonis testing, respectively. Stacked bar plots depicting microbiota composition at bacterial phylum (**N**) and top 10 most abundant genera (**O**), as relative proportion of total 16S V4 amplicon reads within each taxonomic rank.

**Table 1:**
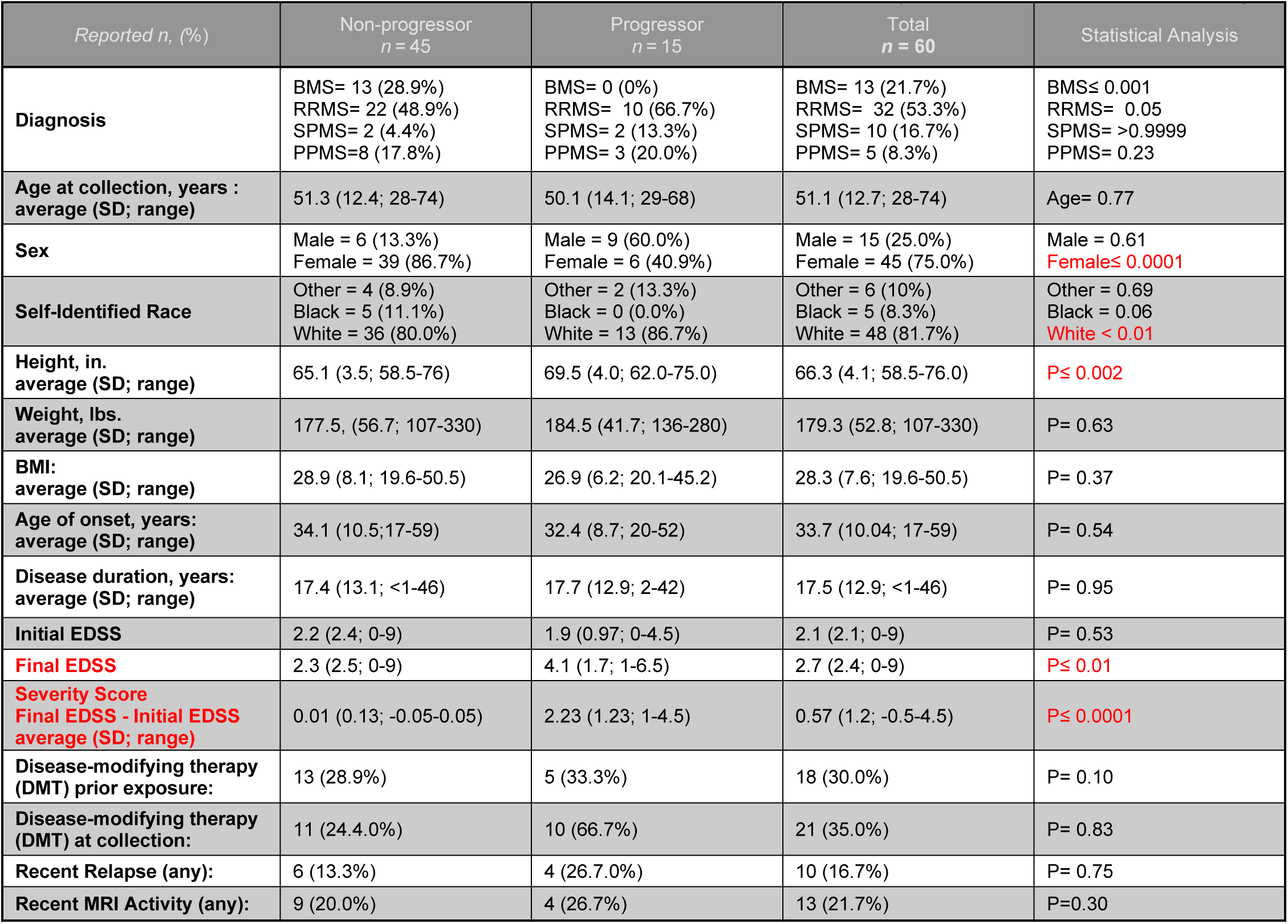
Demographics and clinical characteristics of study participants.

(**Table 1**). To test the hypothesis that the gut microbiome may be an environmental risk factor for predicting disease progression in all MS patients, we grouped patients independent of MS-subtype, based instead on disease progression within the study time period. Specifically, progressor severity was calculated as the change in EDSS from baseline (final EDSS - initial EDSS). Subjects were stratified by progressor severity where a change in EDSS of >|0.5| was considered disease progression (Progressor (P), n = 15), while a change of ≤|0.5| was considered no change in disease severity (Non-Progressor (NP), n = 45) (**Fig. 1B** and **Table 1**). While no differences in initial EDSS between non-progressors and progressors (mean NP=2.2 and P=1.9, p=0.53) was observed, final EDSS was significantly different (mean NP=2.3 and P=4.1, p≤0.01) (**Fig. 1C**, **1D** and **Table 1**). Further, there was the expected difference in MS-subtype between progressor and non-progressors, including an increase in BMS patients as non-progressor (p≤0.001) (**Fig. 1E** and **Table 1**), and a greater proportion of females in non-progressors (p≤0.0001) (**Fig. 1F** and **Table 1**). Importantly, there was a modest but significant overrepresentation of White/Caucasian self-identified patients in progressors (p≤0.01) while no difference in age, BMI, or DMT exposure were observed (**Fig. 1G-K** and **Table 1**).

To pinpoint gut microbial signatures that predispose individuals towards disease progression, we profiled the gut microbiome composition present in baseline fecal samples via 16S rRNA gene sequencing (as detailed in the Methods section). Briefly, DNA was extracted from fecal samples, followed by amplification of the V4 region of the 16S rRNA gene and sequenced using the Illumina MiSeq platform. Reads were trimmed and filtered to remove low quality or short reads, with standard error learning and merging parameters [44]. Following chimera removal (consensus method), an average of 158104 reads per sample remained, with a 77.13% read retention. Amplicon sequence variant (ASV) level taxonomic assignment was performed using the SILVA database (version 138.1) and the established DADA2 (version 1.24.0) pipeline [44–47] resulting in a total 3434 unique ASVs. Downstream data analysis and visualization primarily utilized *phyloseq* (version 1.40.0) with a detailed description of bioinformatic approaches provided in the Materials and Methods section [48].

We first examined gut microbiome diversity between non-progressors and progressors. Consistent with previous studies comparing healthy controls to MS subjects, we found no overt difference in intra-group species richness as measured by alpha diversity (Fisher, Shannon, Simpson, Wilcoxon rank sum tests at p=0.23, p=0.37, and p=0.43 respectively) (**Fig. 1L**) [36, 49]. To examine differences in overall community structure, beta diversity was analyzed between disease progressors and non-progressors. We found no overt difference in community structure as measured by beta diversity (Bray, Unifrac, weighted Unifrac, Wilcoxon rank sum tests at p=0.23, p=0.33, and p=0.43 respectively) (**Fig. 1M**). Taken together, these data suggest that no major changes exist in species diversity within (alpha) or between (beta) subjects that do or do not progress in their disease course. To broadly visualize the composition of the gut microbiome between disease progressors and non-progressor, we examined the distribution of bacterial phyla and genera (**Fig. 1N-0** and **S1**). Bacterial phyla were comprised of the expected taxonomic constituents present within the human gut, including a prominent abundance of *Firmicutes* and *Bacteroidota* (**Fig. 1N**) with no obvious differences between subject groups. Notably, within the top 10 most abundant genera, a contraction in *Akkermansia* and an increase in *Prevotella-9* were visually apparent (**Fig. 1O**), warranting further statistical analysis.

### MS-disease progression is characterized by a unique gut microbial signature

To specifically assess significant changes in gut microbiome composition occurring at each taxonomic level between non-progressor and progressors, differential abundance analysis was performed using DESeq2 (version 1.36.0) [50]. Consistent with exploratory visualization of taxonomic distributions in **Fig. 1N**, we found no individual phyla to be differentially abundant between progressors and non-progressors. However, at each lower taxonomic level, more taxa were found to have significantly lower abundance in the progressor group compared with non-progressors. A single class was depleted in progressors – *Gammaproteobacteria -* driven by the *Enterobacterales* order (**Fig. 2A and 2B**). Additional order level differences included depletion of *Eubacteriales* driven by *Eubacterium* at the family level, *Actinomycetales* with a consistent reduction of the *Actinomyces* genus, and *Monoglobales* which was also depleted at the rank of family (*Monoglobaceae*) and genus (*Monoglobus*) (**Fig. 2B-D**). Notably, the enrichment of the *Clostridiales* order in progressors was carried through to the taxonomic rank of family with a marked increase in *Clostridiaceae* (**Fig. 2B and 2C**). Analysis of bacterial genera revealed an enrichment in *Prevotella-9* and *Alloprevotella*, with a marked depletion of *Ligilactobacillus*, *Holdemanella*, *Erysipelatoclostridium* and the *Ruminococcaceae* member, *CAG-352*, all driven by the differential abundance of individual ASVs (**Fig. 2D and 2E**). Given that ASVs often fail to resolve to the level of species, taxonomy was collapsed to the lowest taxonomic rank assigned and represented as a taxonomic best-hit for fold-change analysis of sequence variants. Differential abundance of ASVs revealed an increase in two *Rhodospirillales* ASVs, and a reduction in several *Lachnospiraceae*, *Desulfovibrionaceae*, *Oscillospiraceae*, *Ruminococcaceae*, and *Peptostreptococcaceae* ASVs (**Fig. 2E**). As fold-change analysis does not account for phylogenetic relationships, differential abundance testing was also modeled using total sum scaling (TSS) log2 linear regression and represented as a taxonomic association heat tree (**Fig. 2F** and **Table S1**). Interestingly, in addition to identifying increases in *Prevotellaceae* and *Alloprevotella* as observed with direct fold-change analysis, a marked depletion of *Akkermansia* (phylum V*errucomicrobiota*) was also identified and was consistent with exploratory observations of the top 10 genera between progressors and non-progressors (**Fig. 1O**, **2F, and Table S1**). Taken together, these data are indicative of a unique baseline gut microbial signature of MS disease progression that is not readily apparent through community structure and diversity analysis alone.

**Figure 2.**
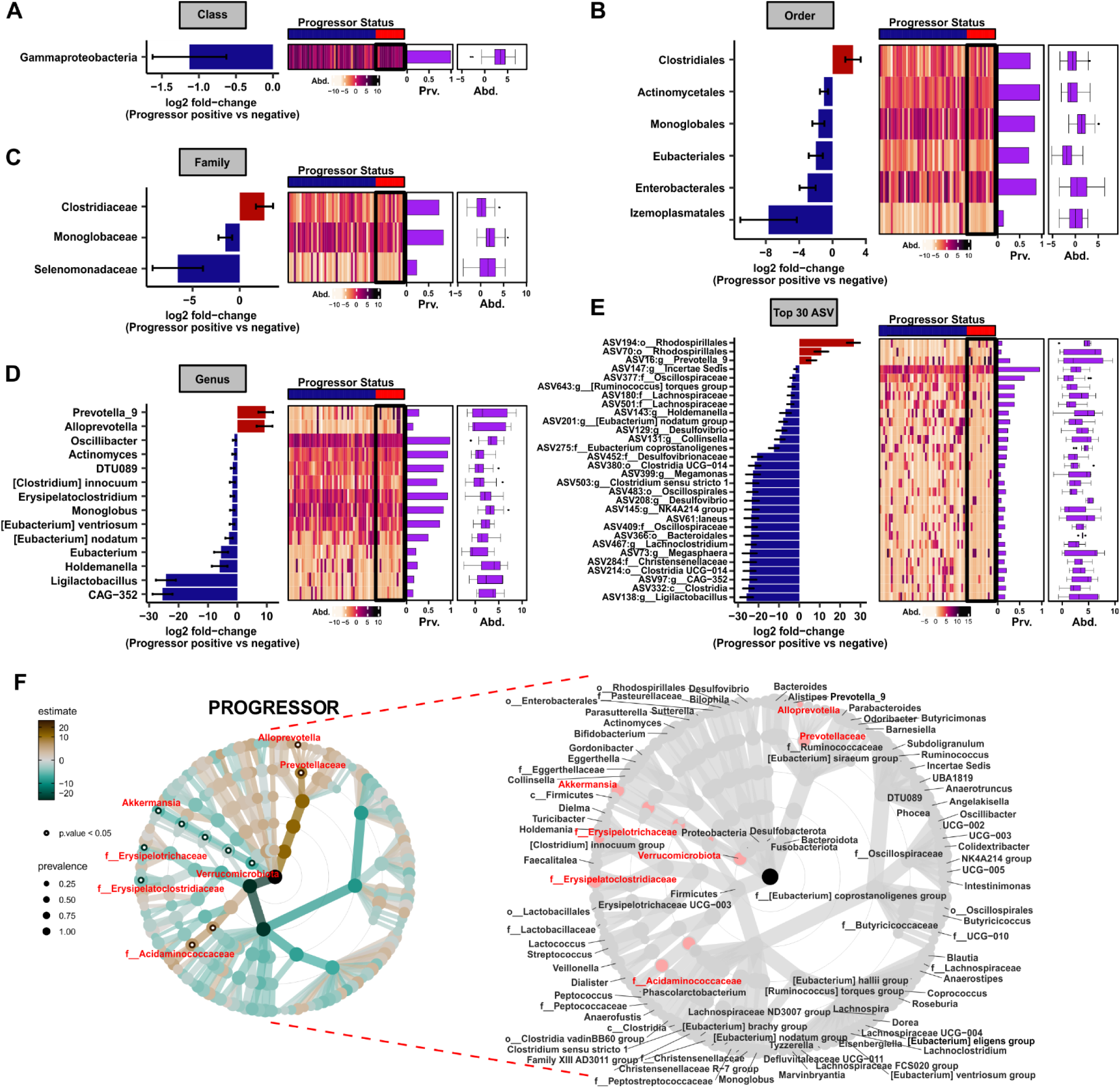
MS disease progressors exhibit a unique baseline gut microbial signature. Differentially abundant taxa between subjects with or without disease progression by (**A**) class, (**B**) order, (**C**) family, (**D**) genus, and (**E**) top 30 ASVs represented as taxonomic best-hit, as determined by DESeq2 analysis, using a cutoff of p_adj_ ≤ 0.05. Log2 fold-change reflects increased abundance in progressors when positive, and decreased abundance when negative. Heatmaps and bar graphs of prevalence as well as center log ratio (CLR) transformed abundance graphs are aligned to the right. (**F**) Taxonomic association heat tree of differentially abundant microbiota determined by total sum scaling (TSS) log2 linear regression through the level of genus. Shown on the right is a phylogenetic key for the heat tree shown on the left. Data was filtered at a minimum prevalence of 0.1, significance determined at a cutoff of p_adj_ ≤ 0.05. Node size indicates proportional prevalence, significant nodes are represented as open circles (or red font in the key), warmer colors are enriched in progressors, with cooler colors indicating depletion.

### Identification of microbiota signatures associated with disease progression

To begin to assess candidate microbiota that most closely correlated specifically with disease progression, gut microbiota collapsed at each taxonomic rank was cross-correlated (Spearman rank correlation) with subject study metadata, focusing on metrics most closely related to disease progression status (progressor or non-progressor), including progressor severity (change in EDSS), initial EDSS, final EDSS, diagnosis/MS-subtype, prior DMT exposure, DMT usage at the time of fecal collection, and sex (**Fig. S2** and **Table S2-S7**). Significantly associated phyla and genera are shown in **Fig. 3A** and **Fig. 3B** respectively. For each taxon strongly associated with progressor status, abundance (center log ratio (CLR) transformed) (**Fig. 3C-3I**), prevalence (ratio of subjects positive for a given taxa) (**Fig. 3J-3P**), and association between progressor severity and microbial abundance (**Fig. 3Q-3W**) were quantified. Consistent with phylogenetic-aware linear regression models (**Fig. 2F**), a reduction in *Verrucomicrobiota* (phylum) and *Akkermansia* (genus) was associated with disease progression (**Fig. 3A-3D, 3J-3K,** and **3Q-3R**), while *Alloprevotella* (genus) was inversely related (**Fig. 3B, 3E, 3L, and 3S**). In addition, an expansion of the *Bacteroidota* phylum was positively associated with both progressor status and severity (**Fig. 3A, 3F, 3M, and 3T**). Novel genera correlating with progressor status included a positive correlation with *Sutterella* and negative correlation with *Defluviitaleaceae UCG-011* and *Lactobacillaceae* as a familial best-hit (**Fig. 3B**, **3G-3I**, **3N-3P**, and **3U-3W)**. Notably, only minor overlap occurred between correlates of DMT exposure and the genera specifically associated with progressor status, suggesting that therapeutics induce unique shifts in the gut flora, and correlations of specific taxa with progressor status are not significantly confounded by treatment status in this patient cohort (**Fig. 3B** and **Table S6**). Interestingly there was also minimal overlap between microbial taxa associated with initial or final EDSS and those associated with progressor status, while *Bacteroidota*, *Akkermansia* and *Alloprevotella* were consistently correlated with progressor severity (change in EDSS) (**Fig. 3A, 3B, Table S2** and **S6**). Abundance, prevalence, and linear regression models of each associated taxon indicated that correlation could be driven by differential abundance alone (*Bacteroidota*, **Fig. 3F**) or both prevalence and microbial abundance (*Verrucomicrobiota*/*Akkermansia*, *Alloprevotella*, *Sutterella*, *Lactobacillaceae*, and *Defluviitaleaceae UCG-011* (**Fig. 3C-3E**, **3G-3I**, **3J-P** and **3N-3P**).

**Figure 3.**
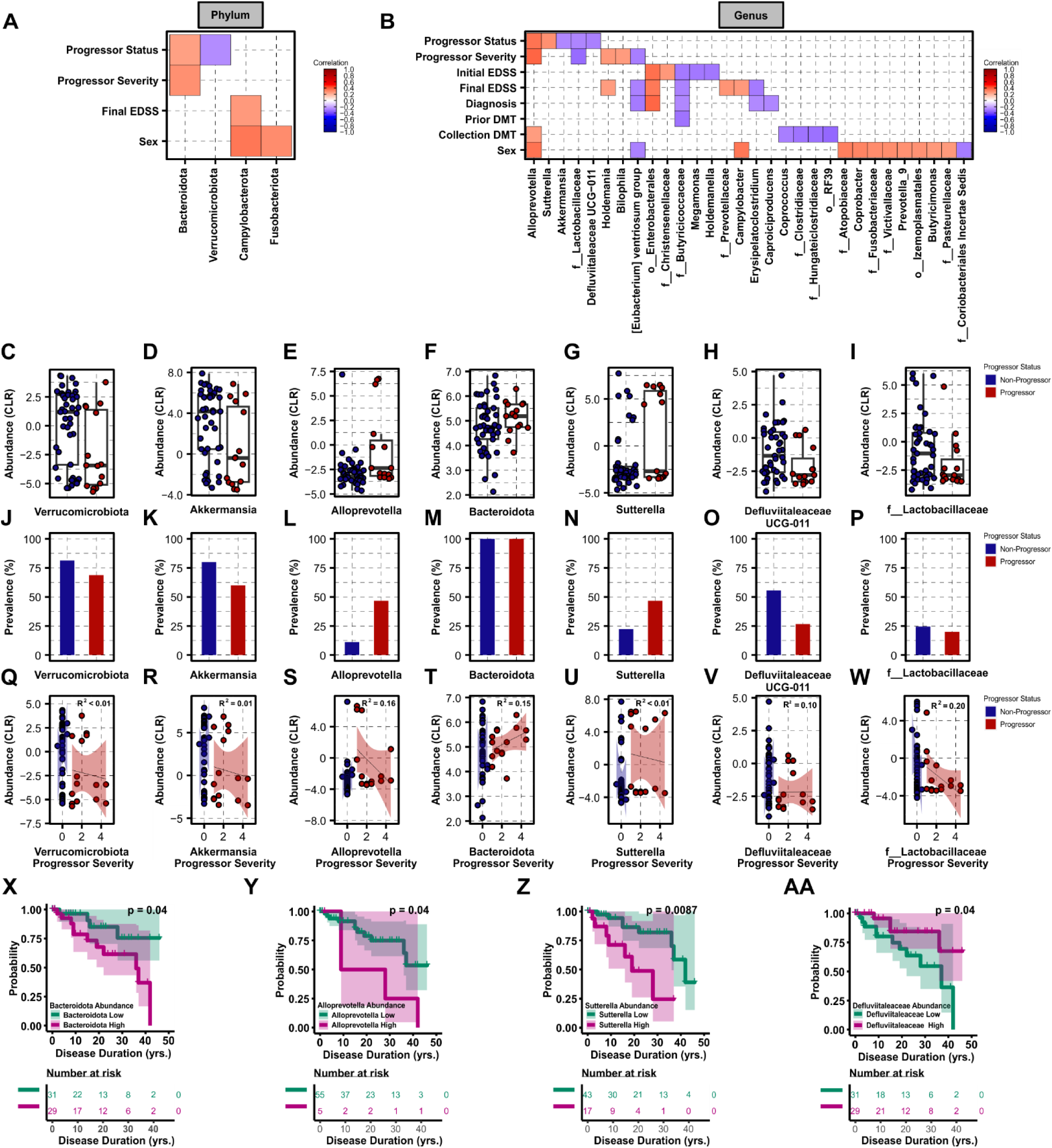
Abundance and prevalence of progressor-associated microbiota. Association of (**A**) phyla and (**B**) genera with progressor status and closely related subject metadata, as determined by Spearman rank correlation ≥|0.2|, at p_adj_ ≤ 0.05. Taxa are sorted from high to low rho-value within each metadata group top to bottom on y-axes, where warmer colors are indicative of positive association (increased abundance) and cooler colors represent negative association (decreased abundance). Metadata binning strategy is listed in **Table 1**. CLR transformed abundance (**C-I**), % prevalence at each taxonomic rank (**J-P**), and linear regression models of progressor severity (change in EDSS over study time period) by CLR transformed abundance (**Q-W**) are shown for each phylum or genera strongly associated with progressor status. Kaplan-Meier curves predicting probability of disease progression over subject disease duration (yrs) using cohorts stratified based on abundance of (**X**) *Bacteroidota* (low/high), (**Y**) *Alloprevotella* (low/high), (**Z**) *Sutterella* (low/high), and (**AA**) *Defluviitaleaceae UCG-011* (low/high). Significance represents log-rank p-value≥0.05 for differential probability of disease progression survival between microbiota driven strata.

To determine if altered microbial abundance of the above identified taxa is sufficient to predict probability of disease progression, median abundance of taxa correlated with progressor status (**Fig. 3A and 3B**) was used to stratify patients (high/low abundance) and Kaplan-Meier analysis was performed. Importantly, binning patients independent of progressor status by low or high abundance of *Bacteroidota* (75.5% (95%CI:49.4%-90.7%) risk of disease progression (HR=3.1 (95%CI:0.98-9.8, p=0.05)) or its genus member *Alloprevotella* (76.3% (95%CI:49.5%-91.4%) risk of disease progression (HR=3.2 (95%CI:0.98-10.57, p=0.05)) was sufficient to significantly stratify patient probability of disease progression over total patient disease duration (log rank p=0.04 and p=0.04, respectively) (**Fig. 3X, 3Y, and S3**), suggesting that *Bacteroidota*/*Alloprevotella* may be useful as a predictive biomarker for future patient disease course. In addition, the genus *Sutterella* abundance, which was positively correlated with progressor status (**Fig. 3B**), was also sufficient to stratify patient probability of disease progression (79.2% risk (95%CI:56.7%-90.4%) with a HR=3.8 (95%CI:1.31-11.13, p=0.01) (**Fig. 3Z** and **S3**). Conversely, *Defluviitaleaceae UCG-011* which was negatively correlated with progressor status, displayed an inverse relationship in predicting disease progression (24.08% risk (95%CI:9.09%-50.3%) (HR=0.32 (95%CI:0.1-1.0, p=0.05) (**Fig. 3AA** and **S3**, log rank p= 0.04). Taken together, these results reveal specific microbial taxa associated with MS disease progression, whose abundance, when taken individually, has some potential to predict disease outcome.

### Vitamin K_2_ and short-chain fatty acid production are depleted in the inferred metagenome of MS progressors

To assess potential functional consequences of the identified changes in microbiota associated with disease progression, total ASVs found to be strongly correlated (Spearman rank correlation ≥|0.2|, p_adj_≤0.05) with progressor status were analyzed using PICRUSt2 [51, 52] to infer their metagenomic functional potential (enzyme commission numbers (E.C.)/designation, KEGG orthology IDS, pathways), followed by differential abundance testing and ASV:E.C.:pathway mapping using MetaCyc [53]. A detailed description of pathway analysis is present in the Materials Methods section and supporting data are provided in supplemental **Tables S7-S12,** with a graphical schematic of analytical approaches provided in **Fig. 4A**. In total 45 ASVs were associated with progressor status, with 19 that were negatively correlated (reduced abundance) and 26 that were positively correlated (increased abundance) in the baseline gut microbiome of patients that went on to exhibit disease progression (**Fig. 4B** and **Table S7**). Progressor status associated ASVs yielded a predicted metagenome encoding 1240 enzymes (**Table S8**) encompassing 253 unique pathways (**Table S9**). Differential abundance testing identified a total of 110 enzymes, 70 of which were predicted to be overrepresented and 40 that were underrepresented (**Fig. 4C** and **Table S10**) within the inferred metagenome of progressor microbiota, comprising 35 differentially abundant pathways, 9 enriched and 36 depleted, respectively (**Fig. 4D** and **Table S11**). Interestingly, a number of pathways predicted to be enriched in the metagenome of progressors were involved in ubiquinol biosynthesis, typically associated with bacterial aerobic respiration, while several depleted pathways suggested a bias towards anaerobic respiration, namely biosynthesis of menaquinol/phylloquinol, the reduced forms of Vitamin K_2_ (**Fig. 4E and 4D**) [54, 55]. Further, baseline progressor gut microbiota were depleted in the capacity to synthesize C2, C3, and C4 short-chain fatty acids (SCFA), including a marked reduction in acetate-production, acetate and succinate fermentation to butanoate, and propanediol degradation to propionate (**Fig. 4D**). To identify putative bacterial species responsible for enriched or depleted enzymes and pathways, the inferred metagenome associated with progressor status was stratified by ASV of origin, and subset to include only ASVs and predicted enzymes statistically associated with progressor status (**Fig. 4E** and **Table S12**). Segregating ASV by EC gene count revealed a total of nine taxa responsible for the top 50 differentially abundant enzymes (**Fig. 4E**), including several ASVs previously shown to be enriched (*Rhodospirillales*, **Fig. 2E**) or depleted (*Akkermansia*, **Fig. 2F**, *Lachnospiraceae* and *Oscillospiraceae*, **Fig. 2E**) in the progressor microbiome. To visualize relationships among select prominent pathways, enzymes, and ASVs, differentially abundant pathways and enzymes were mapped with MetaCyC, manually collapsed at shared nodes, and annotated with progressor status associated ASVs (**Fig. 4F** and **4G**). Interestingly, it was readily apparent that ubiquinol and menaquinol pathways both stem from bacterial chorismate metabolism. Consequently, catabolism of chorismate may therefore represent a critical metabolic branchpoint wherein the over- or under-representation of specific microbiota may favor vitamin K_2_ production (menaquinols) over ubiquinol pathways. Within the ubiquinol superpathways (overrepresented in progressors), a *Rhodospirillales* ASV was predicted to encode two critical enzymes, 2-polyprenyl-6-hydroxyphenol methylase (E.C:2.1.1.222), converting prenyl-benzene-diols into their phenol counterparts, and 3-demethylubiquinol 3-O-methyltransferase (E.C:2.1.1.64), responsible for final ubiquinone production. Interestingly, the same *Rhodospirillales* ASV was also predicted to encode seven copies of glutathione transferase, potentially important for countering bacterial ubiquinone-dependent aerobic metabolism-induced oxidative stress (**Fig. 4E and 4F**). Underrepresentation of menaquinol/phylloquinol pathways in progressors was driven by three ASVs, *A. muciniphila*, a *Coriobacteriia* member (genus DNF00909), and an ASV from the *Veillonella* genus, which were predicted to encode 2-succinyl-6-hydroxy-2,4-cyclohexadiene-1-carboxylate synthase (E.C:4.2.99.20) and o-succinylbenzoate synthase (E.C.:4.2.1.113) involved in 2-succinylbenzoate production from chorismite for downstream menaquinol biosynthesis. To highlight microbiota with a role in restricting SCFA production in MS progressors, the propanediol degradation pathway was also mapped. Both *Oscillospiraceae* and *Lachnospiraceae* (known SCFA producers) were predicted to encode essential enzymes in this pathway and, consistently, their abundance was reduced in the baseline microbiome of progressors (**Fig. 4B, 4E, and 4H**). These data identify candidate species sufficient to induce a bacterial metabolic signature of MS disease progression, which is hallmarked by a reduction in vitamin K_2_ and SCFA production with an increase in aerobic respiration, potentially indicative of elevated oxidative stress within the GI lumen.

**Figure 4.**
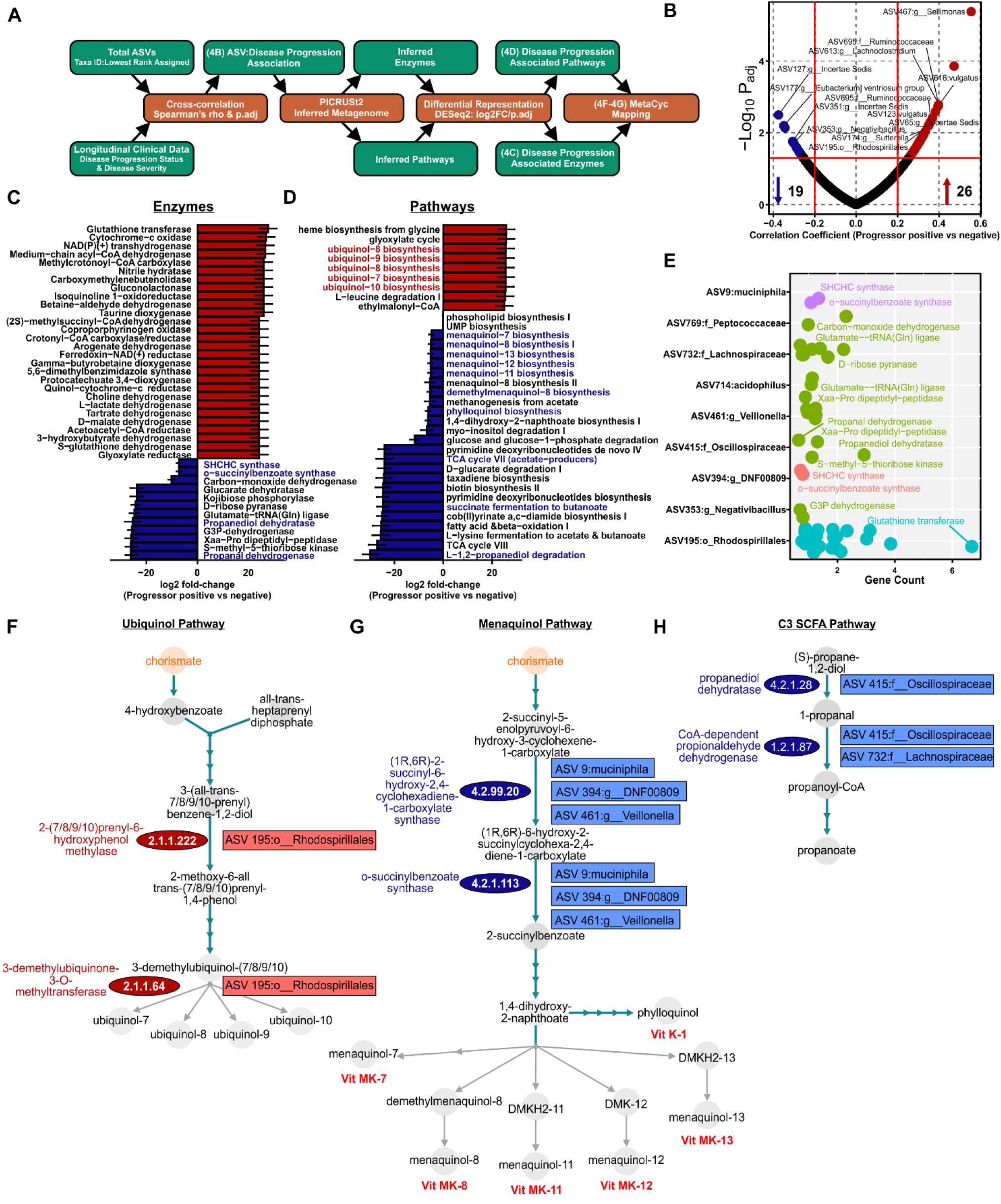
Disease progressors exhibit distinct functional gut microbial metabolic potential. (**A**) Schematic of the computational approach used to infer the functional potential of differentially abundant microbiota associated with disease progression. (**B**) Volcano plot of ASVs (represented as taxonomic best-hits) associated with disease progression as determined by Spearman rank correlation ≥|0.2|, at p_adj_ ≤ 0.05. (**B**) Differentially abundant enzymes and (**C**) pathways in the inferred metagenome of progressor status associated ASVs. Total metagenomic potential was inferred with PICRUSt2 and differential abundance analyzed for the subset of ASVs associated with progressor status using DESeq2 at p_adj_ ≤ 0.05. (**E**) Top 50 differentially abundant enzymes plotted as gene count by ASV (taxonomic best-hit) colored by phylum, as determined by log2 fold-change at p_adj_ ≤ 0.05. Ubiquinol (**F**), menaquinol (**G**), and propanediol degradation (**H**) pathway schematics mapped using MetaCyc and annotated with differentially abundant enzymes and their originating ASVs.

### Baseline gut microbiota are sufficient to predict MS progression

To determine if the baseline gut microbiota and/or clinical features were sufficient to predict disease progression, we leveraged Random Forest-based predictive modeling by training classifiers on: 1) ASV-level total microbiota features, 2) clinical features available at baseline (e.g. excluding final EDSS, progressor status, and progressor severity), 3) ASV-level total microbiota combined with baseline clinical features, 4) restricting microbiota to only progressor status correlated ASVs as identified in **Fig. 4A and 5**) progressor status correlated ASVs combined with clinical features. All classifiers were bootstrapped 500 times using a balanced bagging approach with leave-one-out cross validation. A detailed description of predictive modeling is described in the materials and methods section. Using the first approach with total microbiota ASVs, *A. muciniphila*, *Rhodospirillales*, *Sellimonas*, and *Lachnospiraceae* were identified as contributing most significantly to model performance at a maximum mean decrease in Gini coefficient of 0.24 and mean decrease in accuracy of 2.03 per single ASV (**Fig. 5A**). Receiver operator characteristic (ROC) curve analysis resulted in a moderate area under the curve (AUC) of 0.715. Interestingly, classifiers built on baseline clinical features alone (approach number two), performed similarly to total microbiota, driven by initial EDSS, diagnosis, and collection DMT (mean decrease in Gini coefficient of 1.96, 1.38 and 0.80 respectively) with an AUC of 0.742 upon ROC analysis (**Fig. 5C and 5D**). However, combining ASV-level total microbiota and clinical features (approach number three) failed to further enhance the predictive power of the model over that observed using clinical metadata alone, at an AUC 0.726 (**Fig. S4**), with microbiota features contributing more significantly to overall model performance (**Fig. 5E**). As a fourth approach, to enhance the predictive capacity of the microbiota, classifiers were also trained on just those ASVs identified as associated with progressor status (**Fig. 4B**), which robustly improved model performance resulting in a final AUC of 0.876 in ROC analysis (**Fig. 5F**) and at an out-of-the-box error of 20%, with 7% and 24% class errors for progressors and non-progressors, respectively (**Fig. 5G**). ASVs responsible for the greatest mean decrease in Gini coefficient were consistent with modeling of the total microbiota, including *Akkermansia* and *Rhodospirillales* as the top 2 features of importance (**Fig. 5H**). Consistent with total microbiome trained classifiers, inclusion of clinical metadata along with progressor correlated ASVs (approach number five) did not further enhance classifier performance (**Fig. 5I**). Taken together, these data indicate that the baseline microbiota and patient clinical features are sufficient to predict subsequent patient disease progression, with *Rhodospirillales* and *Akkermansia* contributing most robustly to prediction.

**Figure 5.**
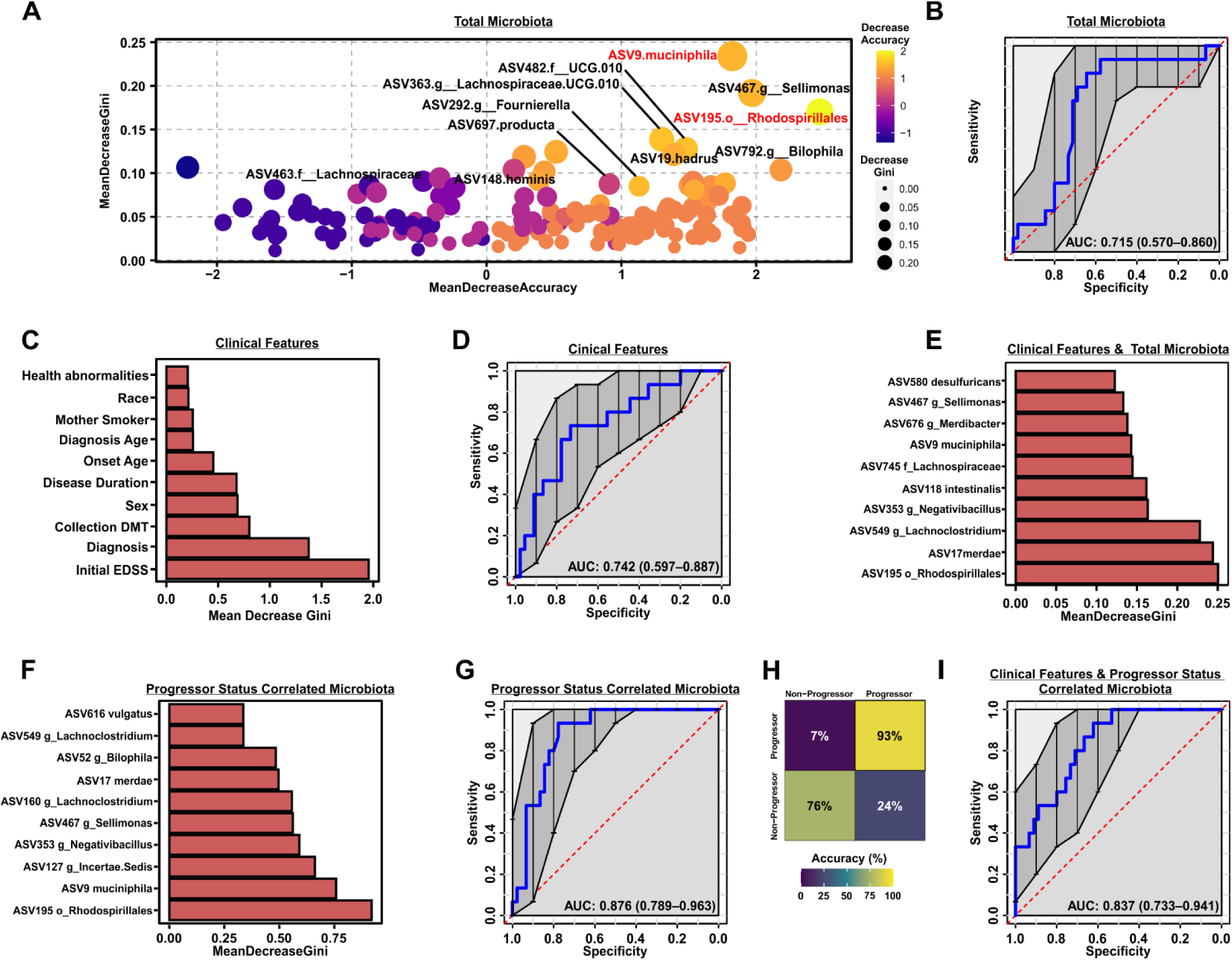
Microbiota composition and patient clinical features are sufficient to predict patient progressor status. (**A**) X-Y scatter plot reflecting total variables of importance by mean decrease in accuracy and mean decrease in Gini coefficient as determined by a Random Forest classifier trained on total ASV level 16S abundance data, with ROC curve analysis shown in (**B**). (**C**) Top 10 clinical features as variables of importance by mean decrease in Gini coefficient as determined by a Random Forest classifier trained on patient metadata available at study baseline, with associated ROC curve shown in (**D**) for clinical data alone and (**E**) when combined with total ASV 16S data. Top 10 variables of importance (**F**), ROC curve (**G**) and class error (**H**) are shown for a Random Forest classifier trained on only the ASVs correlated with progressor status as in Fig. 4B. Inclusion of patient baseline clinical metadata in classifier training of progressor associated microbiota is shown in (**I**). All Random Forest classifiers were bootstrapped 500 times using a balanced bagging approach with leave-one-out cross validation.

### Constipation associated gut microbial changes are modest and independent of progressor status

Detailed metadata on subject demographics, clinical history and any comorbidities was collected from all 60 participants (**Table 1** and **2**). To broadly determine the relationships between each feature, metadata was cross correlated (Spearman rank correlation, ≥|0.2|, p_adj_≤0.05) for hierarchical clustering (Ward.D2 squared minimum variance method) (**Fig. 6A**).Four main feature clusters were observed in metadata correlation, including a small cluster consisting of an intuitive relationship between age and disease duration in years (black border), a second cluster defining relationships between disease progression, disability status and DMT usage (red border), a third cluster segregating recent MRI activity and relapse from age of onset/diagnosis and race/comorbidities (black border) and finally a large central cluster consisting primarily of patient reported symptoms including brain complaints, fatigue, joint or muscle pain (green border). Interestingly, this final cluster also included facets of childhood history (c-section births, breastfeeding and maternal history) as well as allergies (general allergies, food and skin complications). Given that some patient reported metadata features correlated with progressor status, the effect on microbiome structure, as measured by beta diversity, was assessed for each feature as potential confounders. Specifically, the PERMANOVA effect size (as determined by *adonis* R^2^ value) of metadata features associated with weighted UniFrac distance of beta diversity (p_adj_<0.05) was assessed and plotted as a bar graph (**Fig. 6B** and **S6**). Notably only C-section birth passed FDR constraints but had only a modest impact on overall microbial diversity (R^2^=0.04, FDR=0.03) indicating that covariates of progressor status and other patient characteristics do not represent major confounders in this patient cohort.

**Figure 6.**
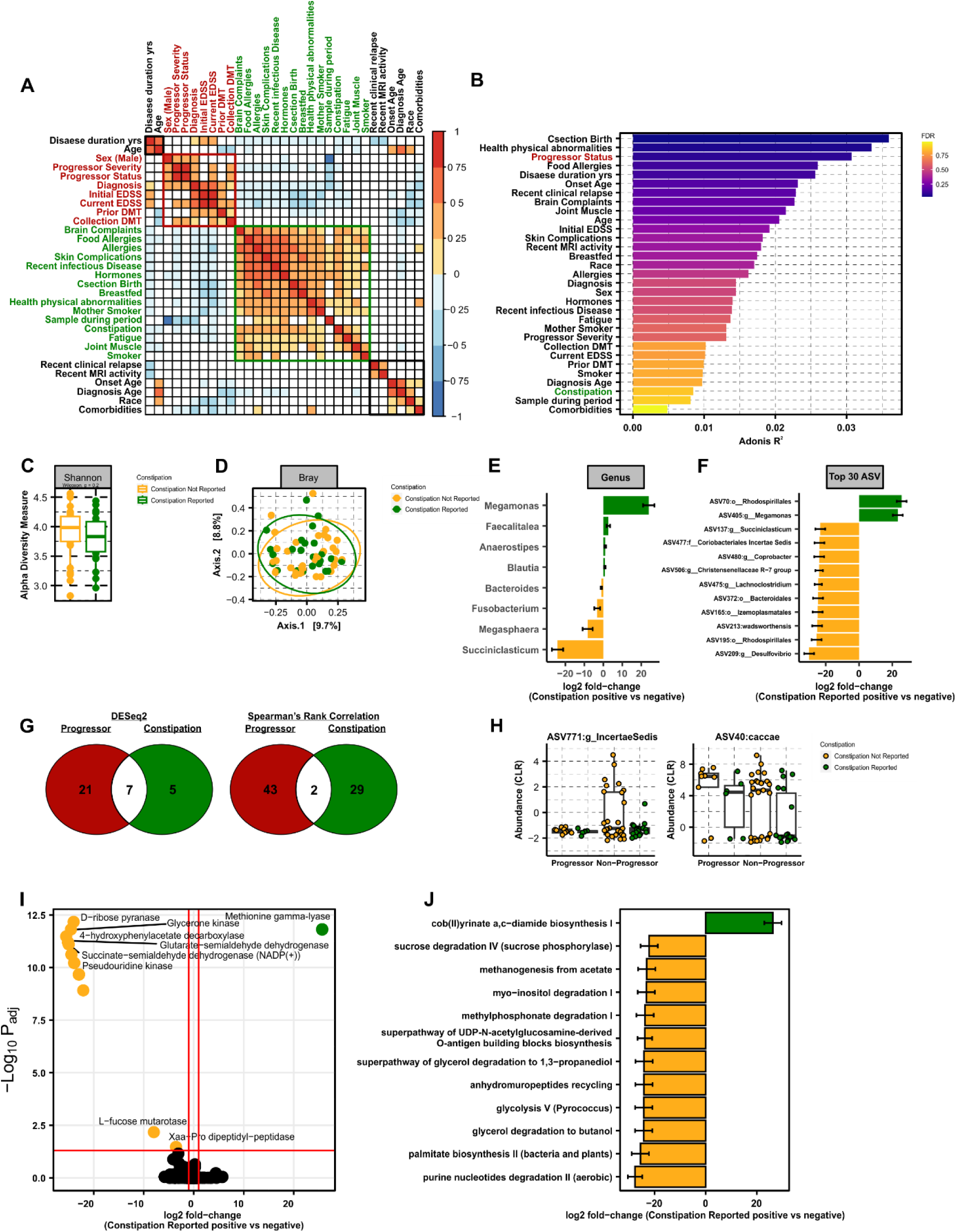
*Constipation is associated with a unique microbial signature that is divergent from that of disease progression*. (**A**) Metadata correlation matrix using a Spearman rank correlation with a significance cutoff at p_adj_≤0.05, where associations not reaching significance are colored in white. (**B**) PERMANOVA of weighted UniFrac distance with the percent of variance explained by each metadata feature as R^2^ on x-axis and colored by FDR, as determined by the *adonis* test. (**C**) Alpha (Shannon) and (**D**) beta (Bray-Curtis dissimilarity) diversity analysis in subjects not reporting or reporting constipation, as analyzed using Wilcoxon rank sum non-parametric or *adonis* tests, respectively. Differentially abundant taxa by (**E**) genus and (**F**) ASVs represented as taxonomic best-hit between subjects with or without constipation, as determined using DEseq2 using a cutoff of p_adj_ ≤ 0.05. Log2 fold-change reflects increased abundance in subjects experiencing constipation when positive and decreased abundance when negative. (**G**) Venn diagrams of shared or divergent ASVs between disease progressors and subjects experiencing constipation as determined by DESeq2 (p_adj_ ≤ 0.05) and Spearman rank correlation ≥|0.2|, at p_adj_ ≤ 0.05. ASVs associated with both progressor status and constipation are plotted as CLR transformed abundance for shared ASVs in (**H**). (**I**) Differentially abundant enzymes and (**J**) pathways from the inferred metagenome of constipation associated ASVs. Total metagenomic potential was inferred with PICRUSt2 and differential abundance analyzed for the subset of ASVs associated with constipation using DESeq2 at p_adj_ ≤ 0.05.

Gastrointestinal (GI) symptoms are frequently suggested to be confounding determinants of MS-specific gut microbial signatures [33, 34]. Consequently, we analyzed the effect of patient reported constipation, as the most frequent GI complaint among MS patients [35], on the gut microbiome, for comparison to alterations associated with progressor status. Consistent with progressor status diversity analysis (**Fig. 1L and 1M**) and permutational analysis of weighted Unifrac (**Fig. 6B**), no overt difference in alpha or beta diversity were driven by constipation (**Fig. 6C and 6D**). Direct fold-change analysis of genera and ASV taxonomic best-hits identified a limited number of taxa that were differentially abundant in patients with constipation, which did not significantly overlap with microbiota that were altered in disease progression (**Fig. 6E-6G**). Specifically, a total eight genera, four increased and four decreased, were associated with constipation, while a total of fourteen differentially abundant genera (two increased and twelve decreased) were observed with progressor status, with no overlap between the two states (**Fig. 6E and 2D**). Strikingly, the abundance of only a total of twelve ASVs (taxonomic best-hit) was altered by constipation (**Fig. 6F**), only seven of which were shared in common with shifts occurring with progressor status (**Fig. 6G**). We previously opted to use Spearman rank correlation ≥|0.2|, p_adj_≤0.05 to predict individual ASVs associated with progressor status prior to inference of predicted metagenomic potential. Comparison of ASV taxonomic best-hits revealed only two shared ASVs associated with both constipation and progressor status (**Fig. 6G-6H** and **Table S7**), including ASV771 in the family *Ruminococcaceae*, genus *Incertae sedis* and ASV40, *Bacteroides caccae* (*B. caccae*). Direct comparison of the abundance profiles of ASVs associated with progressor status and constipation, revealed only ASV771 genus *Incertae sedis* shared the same directionality of change (a moderate decrease) while *B. caccae* abundance was increased in disease progressors but decreased in those experiencing constipation (**Fig. 6H**). To determine if microbial functional profiles rather than individual taxa or ASVs were similar between gastrointestinal symptoms and disease progression, the metagenome of ASVs associated with constipation was inferred using PICRUSt2 for comparison to progressor pathway analysis (**Fig. 4**). In total, 55 enzymes were determined to be differentially represented in the metagenome of subjects experiencing constipation. Of these 55 enzymes, only four were shared with enzymes predicted to be over or underrepresented in disease progressors (EC:5.4.99.62, EC:2.1.1.196, EC:2.1.1.289, EC:3.4.14.11) with only a single enzyme displaying concordant directionality (EC:5.4.99.62, D-ribose pyranase) (**Fig. 6I, 4C**, and **4E**). Interestingly, predicted reduction of D-ribose pyranase was driven by a lower abundance of ASV 714:*Lactobacillus acidophilus* in disease progressors (**Fig. 4E)**, but there was no difference in abundance of this species as associated with constipation (**Fig. S5**). Similar to comparison of differentially represented enzymes, there was minimal overlap in pathway analysis between subjects experiencing constipation and disease progressors (**Fig. 4D, 6J, Table S11**). A total of 12 pathways were altered in the metagenome of patients reporting constipation, with only three pathways shared by progressors including cob(II)yrinate a,c-diamide biosynthesis I (early cobalt insertion), methanogenesis from acetate, and myo-inositol degradation I, with only the latter two exhibiting the same directionality (decreased). These data suggest that constipation is not a major confounder of progressor status associated shifts in the gut microbiome or their inferred functional potential. Moreover, constipation appears to exert a minor impact on the composition of the gut microbiome as compared to MS disease progression, with each displaying a unique microbial and metagenomic signature.

## DISCUSSION

We leveraged longitudinal tracking of disease severity EDSS changes, coupled with baseline assessment of the fecal gut microbiome, in order to identify candidate microbiota predictive of risk of disease progression. Pathway analysis on the inferred metagenome of taxa associated with progression revealed a significant enrichment in oxidative stress-inducing aerobic respiration at the expense of microbial vitamin K2 production, and a decrease in SCFA metabolism. Our study not only showed gut microbial signatures could be used as potential predictors for disease progression, but also linked functional pathways unique to progression. Importantly, only a single previous study leveraged change in EDSS to measure disease progression for correlation with gut microbiome composition, although a metabolic network analysis was not performed [49]. Specifically, Devolder et al. assessed change in EDSS over a 4.4-year period, collecting baseline fecal samples from MS patients. Consistent with our findings, metrics of microbial diversity and community structure were not different between patients who experienced disease progression and those that did not [49]. Additionally, disease worsening was associated with an enterotype replete with *Bacteroides* [49], echoing disease progression associated with the *Bacteroidota* phylum (**Fig. 3A**) in our patient cohort.

The majority of other studies characterizing disease activity-associated microbiota, either compared gut microbiome composition in clinical relapse, between RRMS and progressive forms of disease, or associated microbiota with EDSS at a given cross-sectional timepoint in patient disease course. Thirion et al. reported an elevation in *Coriobacteriia* and *Christensenellaceae* species linked to active disease with relapse (**Fig. 2E** and **Table S1**). Interestingly, a decrease in *Faecalibacterium prausnitzii* (*F. prausnitzii*) was also associated with more disease activity [37], consistent with our results that identified a similar association with an ASV that mapped to the *Faecalibacterium* genus (**Table S7** and **Fig. S7**). BLAST analysis of this ASV revealed 100% coverage and identity to the *F. prausnitzii* 16S rRNA gene. Interestingly, *F. prausnitzii* was also identified as depleted in MS patients in the largest case-control MS microbiome study to date [36]. Cox at al. identified an elevation of *Coriobacteriia* and depletion of *Ruminococcus*, *Oscillibacter*, and *Lachnospiraceae* species associated with worsening disease in RRMS or PMS, consistent with our own analyses of the baseline progressor microbiome (**Fig. 2** and **Fig. 3**) [56]. Interestingly, *A. muciniphila*, as the single member of the *Verrucomicrobia* phylum, was also reported to be depleted in progressive disease, a finding that was also observed in the recent International MS Microbiome Study (iMSMS) and in our own research [36, 56]. Given that *Akkermansia* represents the single bacterial species that is perhaps most consistently reported to be increased in MS patients as compared to healthy controls [18, 19, 36, 57–61], this was an unexpected finding. However, increasing evidence suggests that rather than serving as a disease driver, the host selectively enriches for the abundance of this species, perhaps as a compensatory and/or protective mechanism specific to MS [62]. Our own data identifying a correlation between reduced abundance of *Akkermansia* and progressor status (**Fig. 3A** and **3B**) indeed fits with this model. An alternative explanation may be that *Akkermansia* is increased (in MS relative to the healthy control populations) early during the MS initiation phase, with decreasing abundance upon subsequently worsening disease. How the fluctuation of *Akkermansia* abundance is regulated by the host and any interplay of these mechanisms with MS has yet to be established. However, it is possible that *A. muciniphila* is associated with modulation of mucus production, functioning to restore gut barrier integrity and appropriate tight junction assemblage to prevent dysbiosis [63, 64]. Therefore, a reduction is seen in progressors.

Studies have identified a change in *Prevotella* abundance in the MS gut microbiome, including eight studies reporting a decrease in abundance [18, 19, 27, 40, 60, 65–67]. Consequently, *Prevotella* was tested in preclinical models as a potential probiotic [68, 69]. In contrast, the iMSMS cohort study reported a positive association between several *Prevotella* species and severity of disease in PMS [36], echoing our own data wherein the *Prevotella-9* genus is expanded in disease progressors. Collectively, these data are reminiscent of the above described relationship between *Akkermansia* abundance and disease progression, underscoring the notion that signatures of the gut microbiome in MS may differ from those alterations associated with (or contributing to) disease progression.

No studies have previously documented an increase in the *Rhodospirillales* order in MS. However, recent findings from the International MS Microbiome Study (iMSMS) revealed a correlation between the *Rhodospirillum* genus and cross-sectional disease severity, albeit in opposing directions from our findings, with increased abundance in patients with RRMS and decreased abundance in PMS [36]. In an independent cohort, *Alphaproteobacteria* (class), of which *Rhodospirillium* is a member, was also found to be increased in RRMS [18]. Furthermore, *Rhodospirillaceae* (family) has been linked to Alzheimer’s disease and with a higher risk of irritable bowel syndrome, suggesting a broader association with neurological disease and inflammation [70, 71]. *Rhodospirillales* has also been positively associated with segmented filamentous bacteria, known inflammatory Th17 inducers in the gut, with a posited role in MS pathogenesis [72]. Interestingly, antibiotic treatment in an experimental autoimmune uveitis model resulted in a reduction of *Alphaproteobacteria*, correlating with increased *Gammaproteobacteria*, collectively diminishing the Th17 response and enhancing regulatory T cell proliferation [73]. These findings parallel our own study, wherein *Rhodospirillales* (*Alphaproteobacteria* class) is contracted in non-progressors with a corresponding expansion of *Gammaproteobacteria* (**Fig. 2**).

Though one prior study examined gut microbiome composition in relationship to EDSS changes [49], functional mechanistical understanding of microbiome changes was lacking. We conducted inferred metagenomic analysis and revealed an imbalance between bacterial ubiquinol and menaquinol biosynthetic pathways (**Fig. 4D-4G**). Interestingly, increased ubiquinol at the expense of bacterial menaquinol, as we observed in MS progressors, is a known signature of enhanced bacterial reliance on aerobic respiration in the inflamed gut where oxygen levels are increased [54, 55, 74–79]. Moreover, facultative anaerobes, including *Rhodospirillales* and other *Proteobacteria* phylum members expand under conditions of inflammation and increased luminal oxygen, where they garner a fitness advantage [30, 80–86]. Consistently, *Rhodospirillales*-driven enrichment of 2-(7/8/9/10)prenyl-6-hydroxyphenol methylase (E.C. 2.1.1.222), involved in ubiquinol biosynthesis, was increased in the baseline gut microbiome of patients that would later exhibit disease progression and the single responsible ASV was also predicted to encode glutathione transferase (E.C. 2.5.1.18), necessary to combat oxidative stress induced during aerobic respiration (**Fig. 4E** and **4F**). These data suggest that an early predictor of subsequent MS progression may be inflammation in the gut, triggering increased luminal oxygen availability that favors outgrowth of facultative anaerobes, such as *Rhodospirillales* and other *Proteobacteria*, thus driving overall gut microbiome imbalance.

Increased oxygen-dependent aerobic respiration in the gut has also been linked to a depletion of SCFA-producing obligate anaerobes, mainly including *Clostridial* species, which lack the capacity to combat elevated luminal oxygen [29, 30]. Consistently, we observed a depletion of pathways involved in acetate, propionate, and butyrate production associated with MS progressors and diminished abundance of SCFA-producing microbiota, including *Lactobacillaceae*, *Oscillospiraceae*, and *Lachnospiraceae* members (**Fig. 3E** and **4D**). To highlight the importance of this finding, we identified individual ASVs, depleted in the MS-progressor gut microbiome, that could be causally linked to a reduction in essential genes responsible for propanoate production (**Fig. 4H**). However, dysanaerobiosis, characterized by the increase in oxygen availability within the lumen of the gastrointestinal tract, as a main driver of SCFA-producer reduction, represents a novel emerging mechanism for future study of the MS microbiome, particularly as relevant to disease progression.

In addition to its role in anaerobic respiration for the microbiota, menaquinol biosynthetic pathways are also involved in the production of vitamin K [87]. Interestingly, Lasemi et al. reported strikingly lower levels of vitamin K2 (VK2) in MS patients (235 ± 100 ng/ml vs. 812 ± 154 ng/ml in healthy controls) and that these levels were further reduced in women. Moreover, waning VK2 was linked to relapse and lesions in the CNS [87]. Functionally, vitamin K improves mitochondrial dysfunction [88], inhibits the production of reactive oxygen species to protect neurons and oligodendrocyte precursors from injury [89, 90], and is present at higher concentrations in myelinated regions of the brain, with a role in sphingolipid metabolism [91–94]. Further, there is evidence that vitamin K is beneficial in other neurological diseases, including Parkinson’s [95] and Alzheimer’s disease [96]. In experimental models of MS, Vitamin K increases CNS sulfatides, an essential lipid component of myelin [97], and reduces disease severity when administered prophylactically [98]. Importantly, although a well-known major contributor to bioavailable vitamin K is the gut microbiota, a species-specific link to depletion in MS has not been established. Here, we identified two essential genes involved in vitamin K production contributing to a depletion in menaquinol biosynthesis (**Fig. 4C-4E** and **4G**) that were differentially abundant in the interfered metagenome of patients experiencing disease progression. Moreover, we causally linked these genes to a reduction of three bacterial taxa depleted in the MS-progressor gut microbiome, including, most notably, *Akkermansia*. Consequently, we propose a novel connection between altered *Akkermansia* abundance (one of the most consistent hallmarks of the MS gut) and a reduction in vitamin K as an important area for future research.

To determine if constipation was a major confounder in our own study cohort, we compared the gut microbiome of patients reporting constipation to those exhibiting progression of disease. Strikingly, we observed almost no overlap between taxa associated with progression compared to taxa associated with constipation, with no overlap in the inferred functional potential of differentially associated microbiota. These data suggest that GI dysfunction is not a major contributor to MS progression driven shifts in the microbiota within our study cohort, which is also consistent with approximately equal incidence of constipation in progressors vs. non-progressors (**Table 2**). Interestingly, there are some overlaps between the microbiota associated with constipation here and gut microbiome signatures of MS identified in other patient cohort case-controlled studies. For example, an increase *Blautia* has been observed in four previous studies [18, 38, 41, 66]. Our data suggest that decreased *Blautia* may indicate gastrointestinal dysfunction in MS patients, which is supported by previous work establishing that *Blautia* is also increased with age, BMI, and GI symptoms in MS patients specifically [99, 100]. Similarly, three previous studies [19, 27, 101] reported a decrease in *Sutterella wadsworthensis*, which has also been identified as a treatment-responsive characteristic of constipation [102]. This aligns with our own analysis, which identified a decrease in *S. wadsworthensis* in patients experiencing constipation. Interestingly, several genera exhibited an inverse relationship with known alterations in the MS gut microbiome as compared to constipation-associated shifts highlighted here. For example, both *Megamonas* and [27, 103] *Anaerostipes* [27], are depleted in RRMS, a finding that is in direct opposition to the expansion we observed with constipation. Lastly, *Megasphaera* has been previously associated with GI complaints (in a non-MS cohort), underscoring the specificity in gut microbiome signatures specific to constipation [104]. Taken together, our results identify a distinct microbial signature of constipation in MS patients, which, as highlighted here, can be applied to future and past cross-sectional case-control studies to de-confound comparisons between healthy individuals and people with MS, who frequently report constipation.

**Table 2:** Self-reported symptoms and additional patient history.

| Reported n, (%) | Non-progressor<br>n = 45 | Progressor<br>n = 15 | Total<br>n = 60 | Statistical<br>Analysis |
| --- | --- | --- | --- | --- |
| Constipation reported | 9.0 (20.0%) | 3.0 (20.0%) | 12 (20.0%) | p=0.14 |
| Brain Complaints | 19.0 (42.2%) | 8.0 (53.3%) | 27.0 (45.0%) | p=0.04 |
| Fatigue | 9.0 (20.0%) | 6.0 (40.0%) | 15.0 (25.0%) | p=0.46 |
| Vitamins or dietary supplements | 15.0 (33.3%) | 8.0 (53.3%) | 23.0 (38.3%) | p=0.15 |
| Antibiotic use in the past yr. | 10.0 (22.2%) | 3.0 (20.0%) | 13.0 (21.7%) | p=0.09 |
| Joint or muscle pain | 12.0 (26.7%) | 8.0 (53.3%) | 20.0 (33.3%) | p=0.38 |
| Recent infectious disease | 12.0 (26.7%) | 5.0 (33.3%) | 17.0 (28.3%) | p=0.14 |
| Other reported health conditions | 12.0 (26.7%) | 2.0 (13.3%) | 14.0 (23.3%) | p≤0.01 |
| Sample collected during menstrual cycle | 2.0 (4.4%) | 2.0 (13.3%) | 4.0 (6.7%) | p>0.9999 |
| Hormone use in the past yr. | 8.0 (17.8%) | 2.0 (13.3%) | 10.0 (16.7%) | p=0.11 |
| Current smoker | 9.0 (20.0%) | 3.0 (20.0%) | 12.0 (20.0%) | p=0.14 |
| Biological mother smoked | 13.0 (28.9%) | 5.0 (33.3%) | 18.0 (30.0%) | p=0.14 |
| Born via c-section | 5.0 (11.1%) | 1.0 (6.7%) | 6.0 (10.0%) | p=0.22 |
| Breastfed by mother | 15.0 (33.3%) | 6.0 (40.0%) | 21.0 (35.0%) | p≤0.05 |
| Food allergies or sensitivities | 7.0 (15.6%) | 4.0 (26.7%) | 11.0 (18.3%) | p=0.54 |
| Skin complications | 12.0 (26.7%) | 4.0 (26.7%) | 16.0 (26.7%) | p=0.08 |
| Seasonal or environmental allergies | 19.0 (42.2%) | 5.0 (33.3%) | 24.0 (40.0%) | p≤0.01 |
| Other comorbidity reported | 9.0 (20.0%) | 0.0 (0.0%) | 9.0 (15.0%) | p≤0.01 |

The use of microbiota abundance as a predictive non-invasive and inexpensive biomarker of disease progression is an attractive concept. To determine any predictive value of the microbiota in MS progression, we took two approaches: 1) leveraging association with disease progression to identify of candidate microbiota for subsequent risk assessment using stratification by abundance of a single taxon, and 2) machine learning to identify baseline microbiota signatures of most utility in segregating patients who progress from those who do not. In the first approach, *Bacteroidota*, *Alloprevotella*, *Sutterella*, and *Defluvittaleaceae* abundance was sufficient to stratify patient risk of disease progression (**Fig. 3**). Interestingly, increased *Sutterella* abundance associated with risk of disease progression has been reported previously [67]. However, *Sutterella* levels are also contracted in MS patients as compared to healthy controls [27], and increased abundance has been associated with immunomodulatory treatment intervention, postulated to normalize microbiome composition [19]. Moreover, *Sutterella* is associated with the HLA-DQ8 class II haplotype, which is present at increased frequency in MS and specifically suggested to play a role in disease progression [105–108]. Interestingly, high *Alloprevotella* abundance, associated with disease progression in our study, has also been associated with the progressor DQ8 haplotype [109], though *Alloprevotella* is far less studied in MS specifically. In machine learning approaches, both *Rhodospirillales* and *Akkermansia* were top contributors to predictive models segregating MS-progressors from non-progressors. Predictive modeling restricted to gut microbial composition performed modestly but comparably to the use of readily available patient clinical metadata, including initial EDSS, patient diagnosis, disease duration, and age of onset. Importantly, combining microbiota and clinical features significantly improved predictive performance of disease progression, as did subsetting microbiota features to include only those identified as associated with disease progression (**Fig. 5**). These data serve as proof of principle to demonstrate that analysis gut microbiome composition can be predictive of MS-progression.

MS is a highly heterogeneous disease with various clinical subtypes and accompanying disease courses. Longitudinal tracking of disease progression coupled with baseline analysis of the gut microbiome and patient clinical features offers several distinct advantages including: 1) characterization of disease-relevant microbiome changes, 2) identification of microbiome prognostic makers, 3) offering a built-in internal control, 4) identification of pathological mechanisms, and 5) enhanced translational promise. There is an unmet need to identify patients who are at high risk for progression and need more aggressive therapeutic intervention to prevent long-term disability. Notably, early aggressive medicines potentially pose a greater risk for infection and for other complications compared to traditional therapies; therefore, it is of utmost importance to identify the right patient for aggressive therapy to prevent disability. Our study provides evidence of potentially using the gut microbiome as a prognostic indicator facilitating a more personalized therapeutic approach. Most importantly, our data sheds light on the pathological mechanisms unique to progression, including oxidative stress, Vit K2, and SCFA which could contribute to worsening disease.

## Data Availability

All data produced in the present study are available upon reasonable request to the authors

## Acknowledgements

We thank the University of Michigan Microbiome Core for the Host Microbiome Initiative pilot grant funding of this project. We thank Mr. Michael Dority for his assistance and Drs. Vincent Young, Thomas Schmidt, and Jonathan Golob for consultation.

## METHODS

### Recruitment and study design

A total of 60 MS patients of all types (RRMS, BMS, SPMS, PPMS) were recruited from the University of Michigan (UM) Multiple Sclerosis and the Autoimmunity Center of Excellence (ACE). Patients with BMS were defined as having an EDSS score of ≤3 at least 15 years after disease onset, with no prior MS treatment. As previously described, only true BMS patients who do not have either physical or cognitive impairments were included [13]. RRMS, SPMS and PPMS were defined by the 2010 Revised McDonald criteria [110]. Participant disability status was evaluated at baseline and 4.2 ± 0.97 years later using the expanded disability status scale (EDSS) [111]. Progressors were defined as EDSS progression of >|0.5| modified from that used in the EXPAND trial [112]. All participants had no evidence of relapse or corticosteroid treatment within 3 months prior to baseline and final EDSS assessment. All participants completed a baseline clinical survey reporting subject demographics, disease subtype and history, medications, and any comorbidities as outlined in **Table 1**. No patients reported any dietary restrictions or modifications. Informed consent, approved by the University of Michigan Institutional Review Board, was obtained from patients before study participation. All patients were given written informed consent.

### Specimen collection

Stool was self-collected by subjects using the OMNIgene•GUT (OMR-200) kit (https://www.dnagenotek.com/US/products/collection-microbiome/omnigene-gut/OMR-200.html) and then shipped to UM ACE principal investigator lab within 2 weeks. Stool samples were stored at ambient temperature for less than 4 weeks before DNA extraction.

### Stool sample preparation and 16S DNA sequencing

DNA extraction was performed using the Eppedorf EpMotion liquid handling system following the Qiagen MagAttract PowerMicrobiome kit (Qiagen, catalog no. 27500-4-EP). Sequencing libraries were prepared by the University of Michigan Host Microbiome Core as described previously [113]. Following extraction, samples were quantify using the Quant-iT PicoGreen dsDNA Assay kit (Thermo Fisher, catalog no. P7589). The V4 region of the 16s rRNA gene was amplified from each sample using a dual indexing sequencing strategy in a 20-μl PCR reaction containing 2 μl of 10X Accuprime PCR II buffer (Life Technologies, catalog no. 12346-094), 5 μl of 4 μM universal primers 515-F (5’-GTGCCAGCMGCCGCGGTAA-3’) and reverse 806-R (5’-GGACTACHVGGGTWTCTAAT-3”, 0.15 μl of Accuprime High-Fidelity Polymerase, and 1 μl of DNA template and 11.85 μl of sterile PCR-grade water. PCR cycling conditions were as follows: 2 min at 95°C, 30 cycles at 95°C for 20 s, 55°C for 15 s, and 72°C for 5 min, with a final extension at 72°C for 10 min [114]. PCR products were visualized using an E-Gel 96 with SYBR Safe DNA Gel Stain, 2% (Life technologies, catalog no. G7208-02). Libraries were normalized using SequalPrep Normalization Plate Kit (Life technologies, catalog no. A10510-01), the concentration of the pooled samples determined using Kapa Biosystems Library Quantification kit for Illumina platforms (KapaBiosystems, catalog no. KK4824) and the sizes of the amplicons were determined using the Agilent Bioanalyzer High Sensitivity DNA analysis kit (Agilent, catalog no. 5067-4626). The final library consisted of equal molar amounts, normalized to the pooled plate at the lowest concentration and were prepared according to Illumina’s protocol for preparing libraries for sequencing on the MiSeq (Illumina, catalog no. 15039740 Rev. D) for 2nM libraries. Sequencing was done on the Illumina MiSeq platform, using a MiSeq Reagent Kit V2 500 cycles (Illumina, catalog no. MS102-2003), according to the manufacturer’s instructions with minor modifications [114] including the use of Accuprime High Fidelity Taq (Life Technologies, catalog no. 12346094) in a 5.5 pM reaction to which a 15% PhiX spike was added. Sequencing reagents were prepared according to the Schloss SOP. Custom read 1, read 2 and index primers are added to the reagent cartridge and FASTQ files were generated for paired end reads [114]. Data are available at the NCBI Sequence Read Archive.

### 16S data preprocessing and taxonomic assignment

Raw sequencing reads were analyzed using *DADA2* version 1.24.0 in R version 4.2.1 [44, 115]. Following quality assessment, reads were filtered and trimmed (forward at 240 nt and reverse at 160 nt) and PhiX reads were removed with a maximum number of errors (maxEE) of 2 and quality score of 11. Following standard error learning, forward and reverse reads were merged, and chimeric reads removed using the consensus method. An average of 77.1% of total reads were retained at a mean of 158104 reads per sample. Taxonomy was assigned against the SILVA database (version 138.1) [47]. Unique sequences were aligned using *DECIPHER* version 2.26.0, pairwise distances computed for phylogenetic tree construction with neighbor-joining to fit a maximum likelihood tree using *phangorn* version 2.11.1. The final tree was optimized using a general time-reversible (GTR) model with stochastic rearrangement and rooted using midpoint rooting [116–118]. Taxa, ASV counts, and the final rooted tree were imported into *phyloseq* version 1.40.0 for downstream analysis [48]. Reads that failed to assign at the level of phylum, non-bacterial reads and reads falling below a mean prevalence of 2 where removed, including *Patescibacteria*, *Planctomycetota*, *Spirochaetota*, and *Thermoplasmatota*.

### Diversity analysis

Alpha diversity (Observed, ACE, se.ACE, Shannon, Simpson, InvSimpson, Fisher) [119–123] was calculated using the estimate_richness function in *phyloseq* version 1.40.0 with Wilcoxon rank sum non-parametric testing between groups. Beta diversity (Bray-Curtis, Unifrac, and weighted Unifrac) [124, 125] was calculated using the distance function in *phyloseq* version 1.40.0 and represented as a PCoA of ASV level data with multivariate analysis of variance (adonis2) using the *vegan* package version 2.6-4 [126].

### Taxonomic summaries and differential abundance testing

Closely-related taxa were agglomerated using single-linkage clustering with the tip_glom() function in *phyloseq*. The AGNES algorithm was used with a numeric scalar of 0.2 as the distance threshold for merging taxa [127]. Taxa were also filtered at a 5% prevalence threshold with a mean abundance threshold of 10 reads per sample. Bar graphs represent the top N taxa at each rank as relative abundance of that taxon. The same filtering parameters were applied for differential abundance testing at each taxonomic rank where taxa were agglomerated using the tax_glom function in phyloseq prior to statistical testing using *DESeq2* [50, 128]. Prior to fold-change analysis of individual ASVs, a taxonomic ‘best hit’ was assigned using the format_to_besthit function in the *microbiomeutilities* package version 1.00.16 [129]. Heatmaps and bar graphs of the abundance and prevalence of taxa were generated using centered log ratio (CLR) transformed data with the comp_heatmap function in *microViz* version 0.10.6 [130]. To generate the taxonomic association tree and key, data was normalized using total sum scaling and linear regression was used to model association with progressor status [130, 131]. Model statistics including regression coefficient estimate, test statistics and p-values are contained in **Table S1**. The taxonomic association tree was generated using the taxatree_plots function in *microViz* version 0.10.6 [130].

### Correlation of taxa and clinical features

Following agglomerative clustering consistent with differential abundance testing as outlined above, taxa were filtered at a 3% prevalence threshold with a mean abundance threshold of 10 reads per sample, transformed compositionally (CLR transform), and aggregated at each taxonomic rank. Spearman’s rank correlation of clinical metadata and microbiota was performed using the associate function in the *microbiome* package version 1.18.0 [132] and filtered at rho ≥|0.2|, at p_adj_ ≤ 0.05. Abundance and prevalence of correlated phyla and genera are represented as center log ratio transformed and proportional data, respectively. Linear regression modeling of progressor correlated taxa was performed using the *ggpmisc* package version 0.5.2 [133]. Spearman’s rank correlation of clinical features alone used the *stats* package version 4.2.1 [115] and was plotted using *ggcorrplot* version 0.1.4 with a significant cut-off of p_adj_ ≤ 0.05 and Ward.D2 clustering [134].

### Kaplan-Meier curves

Median abundance of each progressor correlated taxa was used to bin study participants into high or low abundance groups, independent of progressor status. Probability of disease progression over total disease duration in years stratified by low or high abundance of each taxa of interest was estimated using *survminer* version 0.4.9 and *survival* version 3.5-5 and plotted with *ggsurvfit* version 0.3.0 [135–137]. To calculate hazard ratios and confidence intervals, cox regression analysis was performed using *finalfit* version 1.0.6 [138].

### Pathway analysis

To ascertain the inferred metagenomic signature associated specifically with progressor status or constipation, 16S data was subset to include only those ASVs associated with worsening disease or constipation by Spearman’s rank correlation filtered at rho ≥|0.2|, at p_adj_ ≤ 0.05 (**Table S2-7**) and analyzed with *PICRUSt2* to impute functional potential [51, 52]. The resulting per sample unstratified enzyme commission (EC) numbers and MetaCyc pathways including description, were analyzed using DESeq2 for differential abundance testing [53]. ASV stratified matrices of EC numbers were also generated with *PICRUSt2* for mapping of per ASV contribution to enzymes and pathways. Differentially abundant pathways and ECs were imported to MetaCyc as SmartTables to visually represent overlap between pathways, ECs and contributing ASVs.

### Random-forest predictive modeling

Classifiers were generated using the *randomForest* package version 4.7-1.1 [139]. All classifiers used a balanced bagging approach selecting 50% of samples from each class for each tree with class weight balanced by n_total_ / (2*n_class_) and leave one out cross-validation. The optimal number of variables sampled in each node split (mtry), size of terminal nodes (nodesize), and number of trees (ntree) were determined using the trainControl function in *caret* version 6.0-94 for algorithm tuning [140]. Classifiers incorporating clinical features as predictors exclude progressor status, progressor severity, final EDSS and sample during period given this variable is dependent on patient sex. Confusion matrices were plotted using the plot_rf_confusion tool from *modelmisc* version 0.1.1 [141]. Model performance was evaluated as class probability using the predict function in *randomForest*. ROC curves were generated using *ROCR* version 1.0-11 and *pROC* version 1.58.0 [142, 143].

